# The Impact of COVID-19 Non-Pharmaceutical Interventions on Notifiable Infectious Diseases in Poland: A Comprehensive Analysis from 2014-2022

**DOI:** 10.1101/2025.03.05.25323398

**Authors:** Abdulla Hourani, Abdelrahman Abdelsalam, Arman David Sürmeli

## Abstract

**Introduction:** The COVID-19 pandemic prompted widespread implementation of non-pharmaceutical interventions (NPIs) to limit SARS-CoV-2 transmission. These interventions, including lockdowns, face covering, movement restrictions, and school closures, also altered circulation patterns of other pathogens. These measures were enforced on a large scale in Poland beginning in 2020 and persisted through 2022, introducing a rare opportunity to assess broader impacts on other communicable diseases. Previous research frequently addressed short-term alterations in disease incidence, yet knowledge of extended effects remains limited.

**Aim:** The study aimed to assess how the pandemic and associated measures changed the incidence of 17 notifiable infectious diseases in Poland from January 2014 to December 2022. The analysis investigated whether NPIs were correlated with immediate declines, sustained decreases, or rebounds in disease activity, with particular attention to changes in disease dynamics across pre-pandemic and pandemic phases, identifying severity of resurgent trends once restrictions were lifted.

**Materials and Methods:** Biweekly case counts for 17 notifiable diseases were collected from the National Institute of Public Health–National Institute of Hygiene, then aggregated into monthly intervals. The study period encompassed five phases: 2014–2018, 2019 (baseline), and each pandemic year (2020, 2021, 2022). Crude annual incidences were calculated and converted to percentage changes, with comparisons between baseline and pandemic periods as well as among individual pandemic years. Seasonality was removed with X13-ARIMA-SEATS, enabling clearer detection of incidence fluctuations. A two-stage negative binomial generalized linear model (GLM) controlled for autocorrelation and potential confounders, producing incidence rate ratios (IRRs) relative to the 2019 reference phase. The analysis evaluated nine NPIs, including school closure, stay-at-home orders, public gathering bans, and an overall stringency index. Spearman correlation coefficients measured associations between the deseasonalized disease time series, COVID-19 case counts, and each of the nine NPIs.

**Results:** Thirteen of the 17 diseases exhibited cumulative declines from 2020 to 2022 when compared to 2014–2019, ranging from 10.48% (syphilis) to 87.63% (whooping cough). The largest single-year drop appeared in 2021 for whooping cough, which showed a 94.56% decrease from 2019. Statistical modeling revealed an IRR of 0.32 (95%CI 0.24–0.42, p<0.001) in 2020 and 0.11 (95%CI 0.08–0.15, p<0.001) in 2021, exhibiting a significant, sustained reduction in incidence. Scarlet fever, chickenpox, and mumps followed a similar pattern, with IRRs persistently below 0.5 throughout at least one pandemic phase. Invasive Streptococcus pneumoniae remained reduced in 2020 (IRR 0.33 [0.24–0.46], p<0.001), yet rebounded in 2022 with an 88.94% rise over 2019. Clostridium difficile diverged from most other diseases, showing a 2.88% increase in 2020 and a jump of over 117% in 2021 compared to 2019, alongside an IRR of 1.84 (1.64–2.07, p<0.001). Noteworthy surges in norovirus (84.5% in 2021 vs. 2019), HIV (63.5% in 2022 vs. 2019), and syphilis (34.92% in 2022 vs. 2019) aligned with relaxation of NPIs. Correlations generally showed strong negative associations between respiratory pathogens and higher NPI stringency, while C. difficile displayed a positive relationship with COVID-19 case counts and several NPIs. These contrasting trends reflected the multifaceted ways that reduced mobility, physical distancing, and masking influenced various modes of disease transmission.

**Conclusion:** Long-term observation confirmed that widespread NPIs had a strong suppressive effect on many communicable diseases beyond SARS-CoV-2, particularly those transmitted via respiratory droplets. Several infections rebounded when NPIs were relaxed, indicating potential shifts in susceptibility within the population. Future strategies aiming to balance public health protection with social and economic priorities may benefit from these findings, although additional research is needed to clarify how evolving interventions and changing pathogen transmission patterns influence disease resurgence over extended timeframes.

## Introduction

The COVID-19 pandemic, caused by the SARS-CoV-2 virus, has significantly affected both the world’s population and healthcare systems. The rapid global spread of the virus resulted in widespread transmission—a high incidence of cases reaching up to almost 800 million cases, with almost 7 million deaths reported worldwide as of December 23, 2023 [1–4]. In an effort to stop the spread of the virus, nations responded to the pandemic by enacting non-pharmaceutical interventions (NPIs), including lockdowns, social isolation policies, isolating patients with COVID-19, conducting contact tracing, enforcing travel restrictions, cancelling mass gatherings, face covering and school closures [5,6]. The aforementioned measures had impact on halting not only the transmission of COVID-19, but other diseases sharing the same mode of infection or the mode that was disrupted along the used lockdowns (eg. travelling) [7–13]. This situation provides a unique opportunity to investigate the impact of these interventions on the incidence of notifiable infectious diseases and can inform future strategies in infection control.

The first confirmed COVID-19 case in Poland was on the 4th of March 2020 [14] after which cases started to increase drastically [15]. The Polish government implemented multiple NPIs [16], starting with a national lockdown and closure of non-essential businesses. Similar to observations in other nations, it is probable that the (NPIs) implemented in Poland to control the spread of SARS-CoV-2 have led to a decrease in the transmission of other endemic notifiable diseases.

Several studies have investigated the effects of non-pharmaceutical interventions (NPIs) on notifiable infections during the COVID-19 pandemic [7–11]. However, most of these studies focused solely on the outbreak year 2020. While NPIs continued for years, adjusting to the pandemic’s dynamics, their long-term influence on infectious diseases in Poland remains unclear. Therefore, this study aims to address this gap by examining changes in 17 notifiable infectious diseases in Poland before, during, and after the COVID-19 period, spanning from January 1, 2014, to December 31, 2022, using a rigorous statistical methodology.

## Methodology

### Data collection

Biweekly reported cases of 17 notifiable infectious diseases in Poland from January 1, 2014, to December 31, 2022, were collected from the National Institute of Public Health National Institute of Hygiene - National Research Institute (NIPH NIH-NRI) [15]. Diseases with an average of >1000 cases per year between 2014 and 2019 were included: flu & suspicion of flu, whooping cough, scarlet fever, chickenpox, mumps, invasive S. pneumoniae, Salmonella infections, Clostridium difficile enteritis, norovirus enteritis, rotavirus enteritis, giardiasis, infectious diarrhoea & gastroenteritis (Not otherwise specified), hepatitis A, human immunodeficiency virus (HIV), syphilis, Lyme disease, and erysipelas. Hepatitis B and C were excluded due to changes in definitions in 2014 and 2019, respectively. Selected diseases were further grouped into five categories: respiratory and air droplet diseases; gastrointestinal diseases; sexually transmitted and bloodborne diseases; vector-borne diseases; and wound-borne diseases.

The epidemiological surveillance system publishes reports on notifiable infectious diseases biweekly in their public domain (https://www.old.pzh.gov.pl/oldpage/epimeld/). The system is based on electronically gathered, validated, analysed, and interpreted data pertaining to infections and infectious diseases. Physicians are mandated to notify local sanitary inspectors of cases involving communicable diseases. These reports are carefully reviewed by the inspectorates for completeness. Subsequently, the compiled data, along with the case-based questionnaires are forwarded to the regional sanitary inspectorates. The regional inspectorates, in turn, transmit this information to the NIPH NIH-NRI. Following thorough verification and analysis, the NIPH NIH-NRI releases national-level data through periodic biweekly reports [17,18]. Disease definitions are highly consistent with the standards set by the European Centre for Disease Prevention and Control, elucidated comprehensively herein.[19]

The indicators of government restrictions measures were extracted from the Oxford Government Covid Response Tracker (OxCGRT) [16]. OxCGRT was designed by Oxford University’s Blavatnik School of Government to track policy responses in more than 180 countries, rigorously and consistently from January 1, 2020. Data is collected by a research team at Oxford University from publicly available sources, such as government press releases and briefings, international organisation reports, and trusted news articles [20].

Nine non-pharmaceutical interventions were selected, which were: school closure, public events policy, public gathering policy, stay-at-home policy, face covering, public campaigns, public transport policy, internal movement, and international travel policy. The strictness of the NPIs increases with the score of indicators which varies from 0-2 to 0-4. The stringency index, a composite measure based on nine NPIs representing the overall strictness, with a value range of 0 to 100, with 100 being the strictest, was also extracted.

### Statistical Analysis

#### Percentage change

The timeline of the 17 diseases from 2014 to 2022 was divided into 5 phases. Phase I (2014-2018, pre-pandemic), Phase II (2019, the year before the pandemic), Phase III (2020, 1st year of the COVID-19 pandemic), Phase IV (2021, 2nd year of COVID-19 pandemic), and Phase V (2022, 3rd year of COVID-19 pandemic). First, the crude average annual incidence for each disease was calculated with the equation:

*Total cases (year) / Population (year) * 100,000*

Further, the percentage change was calculated between different phases (Table 1) (Graph 1) using the formula:

**Table 1.** The percentage change of the crude average annual incidence of selected notifiable diseases before and after COVID-19.

| Disease | Percent change % |  |  |  |  |  |  |  |
| --- | --- | --- | --- | --- | --- | --- | --- | --- |
|  | 2014- 2018<br>/2019 | 2020<br>/2019 | 2021<br>/2019 | 2022<br>/2019 | 2021<br>/2020 | 2022<br>/2020 | 2022<br>/2021 | 2020-2022<br>/2014-2019 |
| **Respiratory and air-droplet diseases** |  |  |  |  |  |  |  |  |
| Flu or suspicion of Flu | -1.93 | -27.87 | -32 | 3.34 | -5.73 | 43.26 | 51.96 | -18.05 |
| Whooping cough | 10.16 | -77.25 | -94.56 | -89.19 | -76.07 | -52.49 | 98.56 | -87.63 |
| Scarlet fever | 0.03 | -63.58 | -87.33 | -41.96 | -65.21 | 59.38 | 358.18 | -64.3 |
| Chickenpox | -0.33 | -60.02 | -67.6 | -7.42 | -18.97 | 131.58 | 185.79 | -44.92 |
| Mumps | 5.64 | -69.28 | -74.15 | -52.52 | -15.85 | 54.57 | 83.69 | -66.27 |
| Invasive S. pneumonia | -7.78 | -51.01 | -14.15 | 88.94 | 75.23 | 285.66 | 120.09 | 12.29 |
| **Gastrointestinal diseases** |  |  |  |  |  |  |  |  |
| Salmonella | 0.18 | -43.72 | -11.14 | -32.59 | 57.88 | 19.76 | -24.14 | -29.21 |
| C. difficile | -3.25 | 2.88 | 117.32 | 110.98 | 111.24 | 105.09 | -2.92 | 79.98 |
| Giardiasis | 8.16 | -73.47 | -58.09 | -3.88 | 57.96 | 262.3 | 129.36 | -47.3 |
| Rotavirus | -2.9 | -80.01 | -75.05 | 10 | 24.8 | 450.27 | 340.94 | -47.59 |
| Norovirus | -9.01 | -61.87 | 84.5 | 45.87 | 383.94 | 282.61 | -20.94 | 28.63 |
| Diarrhoea | -2.7 | -56.64 | -50.22 | -27.53 | 14.82 | 67.16 | 45.58 | -44.04 |
| Hepatitis A | -2.63 | -88.49 | 4.26 | -76.37 | 805.6 | 105.21 | -77.34 | -52.91 |
| **Sexually transmitted diseases** |  |  |  |  |  |  |  |  |
| HIV | -5.21 | -33.45 | -10.8 | 63.5 | 34.04 | 145.68 | 83.29 | 9.26 |
| Syphilis | -2.83 | -49.65 | -20.5 | 34.92 | 57.9 | 167.97 | 69.71 | -10.48 |
| **Vector- and wound-borne diseases** |  |  |  |  |  |  |  |  |
| Lyme disease | -2.43 | -32.18 | -32.1 | -9.48 | 0.12 | 33.48 | 33.31 | -23.66 |
| Erysipelas | -2.86 | -45.57 | -61.76 | -42.08 | -29.74 | 6.42 | 51.46 | -49.08 |

*(Crude incidence (year) - Crude incidence (reference year) / Crude incidence (reference year) * 100*

Time periods were compared to calculate the aforementioned percentage changes: (average in years of) 2014-2018 and the pandemic years (2020, 2021, 2022) to 2019 as the reference year, the 2nd and 3rd years of the pandemic, 2022 and 2021, to 2020 as the reference year, 2022 to 2021 as the reference year, and lastly, the period of the pandemic (2020-2022) with that of the pre-pandemic (2014-2019).

#### Multivariable generalised linear models

To account for potential confounding factors affecting the NPIs’ effect, such as long-term disease trends, multiple multivariable generalised linear models (GLM) were constructed to quantify the influence of NPIs on each notifiable disease. The phases were defined as defined earlier in percentage change, with Phase II (2019) being the reference. The time-series was deseasonalized using X13-ARIMA-SEATS (Single Extraction in ARIMA time-series) [21,22], and since it operates strictly with monthly and quarterly data, the biweekly reported cases were converted into a monthly format. SEATS is a deseasonalization method that decomposes a time-series into trend, seasonal, transitory, and irregular components. SEATS relies on the foundational assumption that the linearized time series, representing the log of monthly case numbers in the analysis, follows the ARIMA model:

*(B)(BS)(1-B)d(1-Bs)D(yt-xt’) = (B)(Bs)at*

*Where yt is the time series, xt’ is the regression part with covariates xt, at is the white noise with mean 0 and variance 2, and*

*B and Bs: the non-seasonal and seasonal backshift operators, i.e., B(yt) = yt-1, Bs(yt) =yt-12;*

*(B) = 1 − 1B − ⋯ − pBp: a non-seasonal autoregressive (AR) operator of order p;*

*(BS)= 1 − 1BS − ⋯ − PBsP: a seasonal autoregressive operator of order P;*

*(1-B)d and (1-Bs)D: non-seasonal and seasonal differencing operators of orders d and D, respectively;*

*(B) = 1 − 1B − ⋯ − qBq: a non-seasonal moving average (MA) of order q;*

*(Bs) = 1 − 1B − ⋯ − QBSQ: a seasonal moving average of order Q;*

SEATS accounts for the shifts in the mean level of the time series, thus, it can to some extent take the impact of NPIs during the pandemic into account when seasonality is estimated. Two outputs are extracted from SEATS, the deseasonalized monthly cases and the seasonal trend.

A two-stage GLM model using the negative binomial method to account for overdispersion was constructed using the deseasonalized monthly cases from SEATS. In stage 1, the model was fitted with the following predictors: (i) Phase I-V indicators, (ii) long-term trend, (iii) number of person-days (population size times the number of days in the month) as offset. In stage 2, the residuals from stage 1 were extracted and added to the model in stage 1 with a one-month lag to account for autocorrelation. Incidence rate ratios (IRR) of phases 1 and 3-5 estimated by the stage 2 model (Table 2) then reflect the effects of COVID-19 NPIs on the incidence of each selected notifiable disease during that given phase [23,24].

**Table 2.**
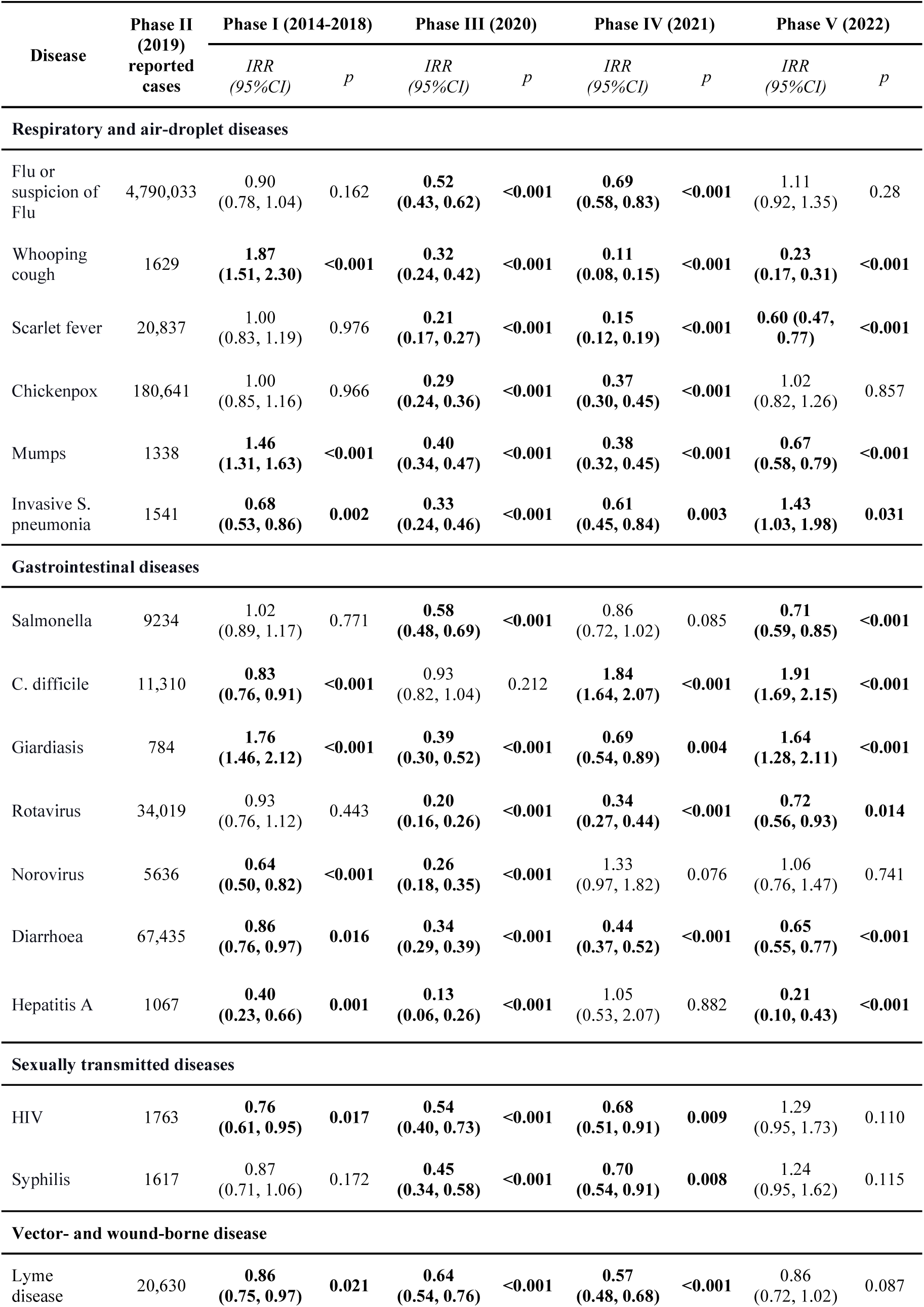
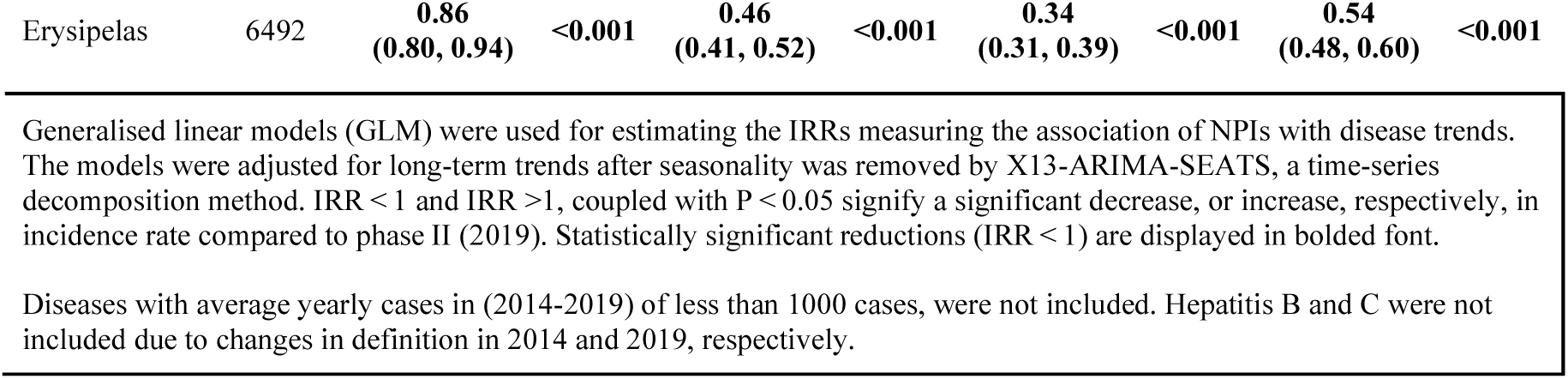
Model estimated incidence rate ratio (IRR) of the selected 17 notifiable diseases.

| Disease | Phase II (2019) reported cases | Phase I (2014-2018) |  | Phase III (2020) |  | Phase IV (2021) |  | Phase V (2022) |  |
| --- | --- | --- | --- | --- | --- | --- | --- | --- | --- |
|  |  | *IRR*<br>*(95%CI)* | *P* | *IRR*<br>*(95%CI)* | *P* | *IRR*<br>*(95%CI)* | *P* | *IRR*<br>*(95%CI)* | *P* |
| Respiratory and air-droplet diseases |  |  |  |  |  |  |  |  |  |
| Flu or suspicion of Flu | 4,790,033 | 0.90<br>(0.78, 1.04) | 0.162 | 0.52<br>(0.43, 0.62) | <0.001 | 0.69<br>(0.58, 0.83) | <0.001 | 1.11<br>(0.92, 1.35) | 0.28 |
| Whooping cough | 1629 | 1.87<br>(1.51, 2.30) | <0.001 | 0.32<br>(0.24, 0.42) | <0.001 | 0.11<br>(0.08, 0.15) | <0.001 | 0.23<br>(0.17, 0.31) | <0.001 |
| Scarlet fever | 20,837 | 1.00<br>(0.83, 1.19) | 0.976 | 0.21<br>(0.17, 0.27) | <0.001 | 0.15<br>(0.12, 0.19) | <0.001 | 0.60 (0.47, 0.77) | <0.001 |
| Chickenpox | 180,641 | 1.00<br>(0.85, 1.16) | 0.966 | 0.29<br>(0.24, 0.36) | <0.001 | 0.37<br>(0.30, 0.45) | <0.001 | 1.02<br>(0.82, 1.26) | 0.857 |
| Mumps | 1338 | 1.46<br>(1.31, 1.63) | <0.001 | 0.40<br>(0.34, 0.47) | <0.001 | 0.38<br>(0.32, 0.45) | <0.001 | 0.67<br>(0.58, 0.79) | <0.001 |
| Invasive S. pneumonia | 1541 | 0.68<br>(0.53, 0.86) | 0.002 | 0.33<br>(0.24, 0.46) | <0.001 | 0.61<br>(0.45, 0.84) | 0.003 | 1.43<br>(1.03, 1.98) | 0.031 |
| Gastrointestinal diseases |  |  |  |  |  |  |  |  |  |
| Salmonella | 9234 | 1.02<br>(0.89, 1.17) | 0.771 | 0.58<br>(0.48, 0.69) | <0.001 | 0.86<br>(0.72, 1.02) | 0.085 | 0.71<br>(0.59, 0.85) | <0.001 |
| C. difficile | 11,310 | 0.83<br>(0.76, 0.91) | <0.001 | 0.93<br>(0.82, 1.04) | 0.212 | 1.84<br>(1.64, 2.07) | <0.001 | 1.91<br>(1.69, 2.15) | <0.001 |
| Giardiasis | 784 | 1.76<br>(1.46, 2.12) | <0.001 | 0.39<br>(0.30, 0.52) | <0.001 | 0.69<br>(0.54, 0.89) | 0.004 | 1.64<br>(1.28, 2.11) | <0.001 |
| Rotavirus | 34,019 | 0.93<br>(0.76, 1.12) | 0.443 | 0.20<br>(0.16, 0.26) | <0.001 | 0.34<br>(0.27, 0.44) | <0.001 | 0.72<br>(0.56, 0.93) | 0.014 |
| Norovirus | 5636 | 0.64<br>(0.50, 0.82) | <0.001 | 0.26<br>(0.18, 0.35) | <0.001 | 1.33<br>(0.97, 1.82) | 0.076 | 1.06<br>(0.76, 1.47) | 0.741 |
| Diarrhoea | 67,435 | 0.86<br>(0.76, 0.97) | 0.016 | 0.34<br>(0.29, 0.39) | <0.001 | 0.44<br>(0.37, 0.52) | <0.001 | 0.65<br>(0.55, 0.77) | <0.001 |
| Hepatitis A | 1067 | 0.40<br>(0.23, 0.66) | 0.001 | 0.13<br>(0.06, 0.26) | <0.001 | 1.05<br>(0.53, 2.07) | 0.882 | 0.21<br>(0.10, 0.43) | <0.001 |
| Sexually transmitted diseases |  |  |  |  |  |  |  |  |  |
| HIV | 1763 | 0.76<br>(0.61, 0.95) | 0.017 | 0.54<br>(0.40, 0.73) | <0.001 | 0.68<br>(0.51, 0.91) | 0.009 | 1.29<br>(0.95, 1.73) | 0.110 |
| Syphilis | 1617 | 0.87<br>(0.71, 1.06) | 0.172 | 0.45<br>(0.34, 0.58) | <0.001 | 0.70<br>(0.54, 0.91) | 0.008 | 1.24<br>(0.95, 1.62) | 0.115 |
| Vector- and wound-borne disease |  |  |  |  |  |  |  |  |  |
| Lyme disease | 20,630 | 0.86<br>(0.75, 0.97) | 0.021 | 0.64<br>(0.54, 0.76) | <0.001 | 0.57<br>(0.48, 0.68) | <0.001 | 0.86<br>(0.72, 1.02) | 0.087 |
| Erysipelas | 6492 | **0.86**<br>**(0.80, 0.94)** | **<0.001** | **0.46**<br>**(0.41, 0.52)** | **<0.001** | **0.34**<br>**(0.31, 0.39)** | **<0.001** | **0.54**<br>**(0.48, 0.60)** | **<0.001** |

Generalised linear models (GLM) were used for estimating the IRRs measuring the association of NPIs with disease trends. The models were adjusted for long-term trends after seasonality was removed by X13-ARIMA-SEATS, a time-series decomposition method. $IRR < 1$ and $IRR > 1$ , coupled with $P < 0.05$ signify a significant decrease, or increase, respectively, in incidence rate compared to phase II (2019). Statistically significant reductions ( $IRR < 1$ ) are displayed in bolded font.
Diseases with average yearly cases in (2014-2019) of less than 1000 cases, were not included. Hepatitis B and C were not included due to changes in definition in 2014 and 2019, respectively.

### Correlation analysis

Correlation analysis was conducted to measure the relationship and its significance between the nine selected NPIs, stringency index, and COVID-19 cases with the deseasonalized 17 selected notifiable diseases (to account for the time-series dependence). Spearman correlation was chosen due to the nature of the data and was done after making sure all the assumptions were met; the correlation coefficients and their associated p-values were extracted (Table 3).

**Table 3.** The correlation coefficient (r) of selected notifiable diseases with the number of COVID-19 case, non-pharmaceutical interventions and stringency index.

| Respiratory and air-droplet diseases |  |  |  |  |  |  |  |
| --- | --- | --- | --- | --- | --- | --- | --- |
|  | Flu & suspicion of Flu | Whooping cough | Scarlet fever | Chickenpox | Mumps | Invasive S. pneumoniae |  |
| Covid-19 | -0.30 | -0.78 | -0.68 | -0.48 | -0.79 | -0.05 |  |
| School Closure | -0.35 | -0.78 | -0.70 | -0.53 | -0.79 | -0.14 |  |
| Public events policy | -0.44 | -0.60 | -0.65 | -0.65 | -0.67 | -0.31 |  |
| Public gathering policy | -0.54 | -0.66 | -0.71 | -0.71 | -0.71 | -0.37 |  |
| Stay at home policy | -0.35 | -0.52 | -0.56 | -0.56 | -0.57 | -0.23 |  |
| Face covering | -0.34 | -0.79 | -0.68 | -0.50 | -0.77 | -0.07 |  |
| Public campaigns | -0.28 | -0.79 | -0.72 | -0.51 | -0.79 | -0.08 |  |
| Public transport policy | -0.45 | -0.40 | -0.50 | -0.50 | -0.50 | -0.46 |  |
| Internal movement | -0.40 | -0.43 | -0.52 | -0.53 | -0.52 | -0.46 |  |
| International travel policy | -0.55 | -0.68 | -0.73 | -0.73 | -0.73 | -0.39 |  |
| Stringency index | -0.35 | -0.78 | -0.72 | -0.58 | -0.82 | -0.15 |  |
| Gastrointestinal diseases |  |  |  |  |  |  |  |
|  | Salmonella | C. difficile | Giardiasis | Rotavirus | Norovirus | Diarrhoea | Hepatitis A |
| Covid-19 | -0.57 | 0.54 | -0.49 | -0.44 | 0.12 | -0.74 | 0.01 |
| School Closure | -0.56 | 0.45 | -0.52 | -0.52 | 0.02 | -0.75 | -0.03 |
| Public events policy | -0.30 | 0.28 | -0.60 | -0.48 | 0.01 | -0.65 | 0.07 |
| Public gathering policy | -0.38 | 0.29 | -0.64 | -0.50 | -0.06 | -0.69 | 0.00 |
| Stay at home policy | -0.18 | 0.34 | -0.49 | -0.47 | 0.10 | -0.56 | 0.18 |
| Face covering | -0.57 | 0.49 | -0.47 | -0.44 | 0.08 | -0.72 | -0.01 |
| Public campaigns | -0.56 | 0.50 | -0.51 | -0.48 | 0.10 | -0.74 | 0.00 |
| Public transport policy | -0.20 | 0.12 | -0.43 | -0.52 | -0.20 | -0.51 | -0.03 |
| Internal movement | -0.17 | 0.12 | **-0.48** | **-0.55** | -0.18 | **-0.54** | -0.02 |
| International travel policy | **-0.42** | **0.24** | **-0.68** | **-0.50** | -0.10 | **-0.72** | -0.06 |
| Stringency index | **-0.54** | **0.47** | **-0.56** | **-0.54** | 0.04 | **-0.77** | 0.00 |

**Sexually transmitted diseases, vector- & wound-borne diseases**
|  | HIV | Syphilis | Lyme disease | Erysipelas |
| --- | --- | --- | --- | --- |
| Covid-19 | -0.03 | **-0.26** | **-0.44** | **-0.79** |
| School Closure | -0.14 | **-0.30** | **-0.43** | **-0.79** |
| Public events policy | **-0.31** | **-0.45** | **-0.47** | **-0.64** |
| Public gathering policy | **-0.36** | **-0.51** | **-0.51** | **-0.69** |
| Stay at home policy | **-0.27** | **-0.31** | **-0.38** | **-0.56** |
| Face covering | -0.05 | **-0.27** | **-0.44** | **-0.79** |
| Public campaigns | -0.08 | **-0.29** | **-0.45** | **-0.81** |
| Public transport policy | **-0.38** | **-0.39** | **-0.26** | **-0.45** |
| Internal movement | **-0.40** | **-0.36** | **-0.30** | **-0.47** |
| International travel policy | **-0.37** | **-0.54** | **-0.54** | **-0.71** |
| Stringency index | -0.14 | **-0.33** | **-0.46** | **-0.80** |

\* $P < 0.05$ are displayed in bolded font
\*\* $P < 0.01$ are displayed in bolded italics font

## Results

### Percentage change of the 17 selected diseases before and after COVID-19

As shown in table 1, compared to the pre-COVID period spanning from 2014 to 2019, 13 out of the 17 chosen diseases experienced a decline in the COVID era (2020-2022), ranging from 10.48% to 87.63%. With the exception of C. difficile, all selected diseases exhibited a decrease in 2020 compared to 2019. Similarly, in Phase IV (2021), a similar trend was noted, except for norovirus and hepatitis A, which saw an increase of 84.5% and 4.26%, respectively. In Phase V (2022), alongside C. difficile and norovirus, other diseases started to rise, namely flu, invasive S. pneumoniae, HIV, and syphilis. Notably, invasive S. pneumoniae and HIV recorded the most substantial increases at 88.94% and 63.50%, respectively.

When comparing intra-pandemic periods, the percentage change for 2021/2020 indicates a decrease in all respiratory and airborne droplet diseases, except for invasive S. pneumoniae, which saw a significant increase of over 75%. Conversely, all gastrointestinal and sexually transmitted diseases experienced substantial increases. Remarkably, hepatitis A showed a dramatic surge of over 800%, followed by norovirus with an almost 384% increase. Lyme disease cases remained relatively stable in both years, whereas there was a substantial increase of almost 30% in erysipelas cases. Interestingly, the percentage changes for 2022/2020 revealed a significant rise in nearly all diseases, ranging from approximately 7% to about 285%. The only exception was whooping cough, which saw a sharp decline of over 50%. For the 2022/2021 percentage change, most diseases exhibited an increase, except for four gastrointestinal infections, namely Salmonella, C. difficile, norovirus, and hepatitis A. The latter of which experienced a notable decrease of over 77%.

### Generalised linear model estimated association of NPIs with disease trend

In delineating the outcomes derived from the 2nd stage of the GLM model, several key patterns emerged, reflecting the nuanced impact of NPIs and the COVID-19 pandemic across different disease categories. The comprehensive investigation spanned four pivotal phases - III to V - and encompassed a spectrum of 17 notifiable diseases, revealing noteworthy shifts in incidence rates. Beginning with the broader context, excluding C. difficile, the IRRs of all diseases displayed a discernible reduction in at least one of the pandemic phases (III-V). Particularly striking was the consistent significant reduction observed in whooping cough, scarlet fever, mumps, rotavirus, diarrhoea & gastroenteritis, and erysipelas across all three phases, suggesting an enduring influence of NPIs on these specific diseases.

As presented in table 2, maximal reduction was observed during Phase III (2020), where 16 out of the 17 selected diseases experienced a significant decrease. Notably, the majority of diseases witnessed the most substantial reduction in incidence during this phase, with exceptions noted for whooping cough, scarlet fever, mumps, Lyme disease, and erysipelas. Intriguingly, the incidence of these diseases exhibited a more pronounced decline in Phase IV (2021). The subsequent phases - 2021 and 2022 - saw diminishing numbers of diseases exhibiting significant reductions (13 out of 17 and 8 out of 17, respectively), indicating a waning impact of NPIs from 2020 to 2022.

The overarching impact of NPIs was most pronounced in respiratory diseases throughout the pandemic. Notably, whooping cough demonstrated a remarkable decline to 0.32 (95%CI 0.24-0.42) in 2020-0.11 (0.08-0.15) in 2021, and 0.23 (0.17-0.31) in 2022. All respiratory diseases exhibited a significant reduction in Phase IV. Conversely, the incidence of invasive S. pneumoniae increased significantly in Phase V (2022) with an IRR of 1.43 (1.03-1.98. For gastrointestinal diseases, both diarrhoea & gastroenteritis and norovirus displayed a steady decline in incidence throughout the pandemic. This trend does not stay consistent, such as in the case of C. difficile, which exhibited a remarkable increase in incidence during Phases III (IRR: 1.84, 1.64-2.07) and IV (IRR: 1.91, 1.69-2.15), and in the incidence of giardiasis, also experiencing a significant increase in Phase IV (IRR: 1.64,: 1.28-2.11). A clear reduction in the prevalence of sexually transmitted diseases (STDs) was observed during Phase III, succeeded by a more moderate decline in Phase IV, with incidence levels exhibiting relative stability in Phase V.

### Correlation analysis

The monthly number of COVID-19 cases demonstrated a negative correlation with the majority of selected diseases, with the highest negative correlation being with mumps (r=-0.79) and erysipelas (r=-0.79). Notably, hepatitis A exhibited no significant correlation with either NPIs or COVID-19, norovirus only showed a negative correlation with the public transport policy (r = −0.2), and C. difficile displayed a notable positive correlation with almost all NPIs and COVID-19.

For the remaining diseases, school closure, face covering, and public campaigns were consistently negatively correlated with all diseases except for invasive S. pneumoniae and HIV, where no significant correlation was observed. Stay-at-home policy and internal movement were concurrently negatively associated with all remaining diseases except for Salmonella infections, which exhibited an insignificant correlation. Public events policy, public gathering policy, public transport policy, and international travel policy were all negatively associated with the remaining 14 diseases. The highest correlation was observed with mumps (r=-0.82), with nearly all diseases showing a significant negative correlation with NPIs except for 5, namely, invasive S. pneumoniae, C. difficile, norovirus, hepatitis A, and HIV. (see Table 3)

## Discussion

Through the use of publicly available data from the Polish Information System for Disease Control and Prevention and analysis of pre-, intra-, and post-pandemic periods, the present study effectively quantifies the effect of NPIs on the prevalence of notifiable diseases. The distinction of phases and subsequent comparison between given time periods aids in the isolation of the spread and impact of specific disease, not only through the trends of said disease and whether they declined but also through a pattern of how NPIs influenced this trend. The practical application of NPIs is a very convoluted process, with the stringency of interventions constantly shifting, and although the use of population behaviour and epidemiological tracking helps aid in decision-making [21], the methodological concerns of public policy and therefore public action, in the context of proper application and evidence in correlation between concurrent NPIs is significant, necessitating an observational analysis of correlation rather than pure isolation of any finding. From mandates implemented and their subsequent relaxation, the study uses summarised analysis of the relationship between NPIs and their effectiveness rather than individual policy and how it factored into the specific issue it was attempting to mitigate at the time, since even in the case of the nationwide mask mandate implemented across Poland on October 8, 2020 having increased the OxCGRT scrutiny value to its highest levels of the year [20], it would be difficult to isolate whether the consequences are related to its original justification.

NPIs implemented during the COVID-19 pandemic had a unique quality in how their method impacted the spread of disease, specifically attempting to mitigate human-to-human contact with an emphasis on COVID-19’s respiratory droplet transmission. This resulted in the greatest impacting correlation being related to said methods of transmission. Respiratory and air-droplet-related diseases generally noticed equal or near equal incidence rate ratios (IRR) when comparing pre-pandemic phases to the onset of the pandemic and implementation of NPIs (reference phase: 2019/Phase II), however, significantly dropped as interventions related to decreasing public contact in relation to significant correlation factors of disease were implemented. All recorded notifiable diseases acknowledged in this study received a decrease in IRR (at associated confidence intervals (CI) of 95%) from Phase I to Phase III (Table 2) (except in C. difficile, which will be expanded upon). This baseline was not a continued trend for flu, chickenpox and invasive S. pneumoniae. Flu (Phase I 0.52 (95%CI 0.43-0.62)) exhibited a trend toward normal levels during Phase IV (0.69 (95%CI 0.58-0.83)), which it would end up exhibiting during later years (Phase V 1.11 (95%CI 0.92, 1.35)) after NPIs were eased, although at a much lower significance which is to be noted. With the increased vaccination intra-pandemic and increased availability of diagnostic testing post-pandemic [25,26], there was a noticeable bias on detection rates which would equate to low incidence rates during Phase I-III but a sharp increase with Phase V, also coinciding with a decrease in NPI stringency during this period [20]. Chickenpox (Phase III 0.29 (95%CI 0.24-0.36)) and invasive S. pneumoniae (Phase III 0.33 (95%CI 0.24-0.46)) exhibited similar phase trends with the only difference being the latter’s significance maintaining a lack of deviation and also proving to be at a much harsher consequence (Phase V 1.43 (95%CI 1.03, 1.98)) in reflection to pre-COVID-19 levels. Whooping cough showed the most drastic decreases in incidence and exhibited lasting impacts post-pandemic, at an over 4-fold reduction (Phase V 0.23 (95%CI 0.17-0.31)). With the cyclical nature of whooping cough being every 3 to 5 years, this lasting effect post-pandemic could have been due to external factors delaying the next cyclical resurgence of the virus [27,28]. Scarlet fever (Phase III 0.21 (95%CI 0.17-0.27)) and mumps (Phase III 0.40 (95%CI 0.34-0.47)) both exhibited statistically significant decreases throughout Phases I-IV but did exhibit an increase in Phases IV-V although still at lower than pre-pandemic levels (Phase I 1.00 (95%CI 0.83, 1.19), 1.46 (95%CI 1.31, 1.63) respectively). The changes among incidence suggest potential shifts in disease dynamics amidst changing NPI measures and highlight the effectiveness of NPIs in mitigating respiratory disease transmission. With decrease of incidence of the aforementioned diseases mirroring decreases in cases of COVID-19 [29], independent of NPI stingency, under- or mis-reporting of cases is unlikely. The closure of settings where respiratory disease spreads such as schools and restaurants would decrease their incidence [30], conversely, alleviating stringency would correlate their increase. Interestingly, the same trend can still be seen within non-respiratory pathogens however, such as the fecal-oral route of norovirus not relating to the normative [31] droplet-related respiratory disease mode of transmission [32], alluding to additional benefits of COVID-19 related NPIs to said pathogens.

Other types of pathogens, such as those transmitted through gastrointestinal, sexually transmitted, or vector-borne routes, may have also experienced correlated effects even if respiratory pathogens were arguably the most impacted by NPIs. The starkly influenced shift in human movement and behavior during this period likely altered exposure levels [17,21], by not only limiting public exposure but also through mandates restricting movement, potentially influencing the transmission dynamics of various infectious diseases beyond respiratory ones. Salmonella, diarrhoea & gastroenteritis, and rotavirus all exhibited long-term decreases in IRRs (Phase V 0.71 (95%CI 0.59-0.85,) 0.72 (95%CI 0.56-0.93)-0.65 (95%CI 0.55-0.77) respectively) compared to Phase I (Phase I 1.02 (95%CI 0.89, 1.17)-0.93 (95%CI 0.76-1.12)-0.86 (95%CI 0.76-0.97) respectively). Hepatitis A also showed long-term decreases (Phase I 0.40 (95%CI 0.23-0.66) to Phase V 0.21 (95%CI 0.10-0.43)) in incidence, but this was seemingly due to being highly prevalent or reported in Phase II, skewing all significant IRRs (Phase III 0.13 (95%CI 0.06-0.26) and omitting Phase IV) in the observed time frame to lower values. Giardiasis and norovirus showed minimal change, with a slight decrease or increase in IRR when comparing Phase I to V (Giardiasis: 1.76 (95%CI 1.46, 2.12) to 1.64 (95%CI 1.28, 2.11), norovirus: 0.64 (95%CI 0.50-0.82) to 1.06 (95%CI 0.76, 1.47) (with the greater absolute change in norovirus being attributed to its low statistical significance)), although exhibiting the same pattern of decrease during Phase III with a followed gradual increase to normative values. Lastly, C. difficile displayed a positive correlation with all NPIs and also was the only of the 17 studied diseases to not show the same trend, namely decreasing during the 2020/2019 (Phase III) periods (Phase I 0.83 (95%CI 0.76-0.91), Phase III 0.93 (95%CI 0.82, 1.04). C. difficile infections have been corroborated to increase mortality rates in COVID-19 patients [33,34] assumingly to have had a change in incidence due to gut dysbiosis and poor antibiotic stewardship exacerbating the concurrent infection and impacting patient outcome [33]. However, these results are inconclusive, with literature analysing incidence also corroborating results proving non-statistically significant increase or decrease in C.difficile [35,36] likely due to increased contact isolation even if the [36–38] increased antibiotic use in treating COVID-19 patients would exasperate C.difficile infection rates normally [39].

Sexually transmitted, vector- and wound-borne disease were the least impacted by the studied NPIs, COVID-19, and among the studied diseases (COVID-19 correlation: HIV −0.03, syphilis −0.26, Lyme disease −0.44), with the notable exception of travel restricting NPIs (internal movement: −0.40, −0.36, −0.30 and international travel policy-0.37, −0.54, −0.54), due to its direct impact on the mode of transmission. Erysipelas was an outlier among all of the aforementioned diseases, proving the strongest absolute correlation with all NPIs in its category and with a decrease in IRR over time (Phase I 0.86 (95%CI 0.80-0.94) Phase V 0.54 (95%CI 0.48-0.60)). To note, changes in reporting and tracking of sexually transmitted diseases, specifically syphilis [40] during pandemic levels, might affect the incidence due to incongruencies in reporting rather than a real reflection of trends. This was most evident during the initial prevalence of the omicron variant during January and February of 2022 (Phase V) wherein a surge of migration of war refugees from Ukraine caused issues in tracking and reporting, while the decrease in NPI stringency gave way to a significant increase in the rate of HIV high-risk behaviour in the post-lockdown era, especially among men-who-have-sex-with-men in conjunction to the prevalence of drug abuse in persons-who-inject-drugs [41,42]. With challenges in managing HIV incidence in Ukraine already prevalent prior to the onset of the conflict, only 69% of an estimated 260 thousand HIV positive were aware of their infection status and of those, only 57% were receiving antiretroviral therapy [43], which may have resulted in an increase in reporting of cases among migrants within Poland during the refugee influx when compared to the general population [41]. The immense inflow of migrants, totalling 1.8 million during the first month of the war [44], likely influenced the incidence and epidemiology of sexually transmitted infections during this time, where although the aforementioned testing of sexually transmitted diseases decreased during pandemic levels, the subsequent increase of cases may be isolated to have coincided with these migration events due to the increase of COVID-19 vaccines during 2021, the onset of a clinically less severe SARS-CoV-2 Omicron variant [45], and the decrease in NPI stringency resulting in improved rates in testing of sexually transmitted, vector-and wound-borne disease [17,41,46]. Internal movement and international travel policy changes and easing during this time likely impacted and exacerbated incidence within Poland, making increased efforts to provide (equity in) testing vital in mitigation as evident with a need for optimised diagnosing and tracking of notifiable infectious diseases amongst migrants and refugees [42].

With the implementation of NPIs reducing immune stimulation in a population due to the suppression of circulation of disease, a growing fear is the concept of “immunity debt,” the realisation of pandemic consequences totaling a growing population of increased susceptibility and naive immune systems to pathogens [47]. This declined herd immunity would result in a higher likelihood and severity of future pandemics [48], exasperated by the duration and magnitude of NPIs implemented [47,49]. Further analysis of long-term consequences is required, although this study does exhibit a trend of increase in disease during post-pandemic periods, possibly due to said broad population immunity concerns.

Previous studies have gone in-depth by analysing country-wide and district-specific data, often tackling the same diseases by overlap of countries’ notifiable diseases [7–11,19]. However, this study contrasts with current literature by increasing the scope of the observational timeframe, shifting from a window of 2020 [8,9,11,12] and expanding it to include “post-pandemic” years to bring into perspective the long-term impacts of NPIs with data from 2021 and 2022. As previously mentioned, some diseases did exhibit sustained changes from the inclusion of NPIs and did not simply return to Phase I or Phase II values, creating a more nuanced view on how/what policy shaped the current epidemiological landscape. This addition is compounded by how NPIs were defined in this study; the main difference being that previous research classified NPIs in the context of governmental policy and public health guidelines, while this study focused on applying quantitative indicators to assess impact more objectively. This change results in a more data-driven and conclusive analysis of the relationship between NPIs and disease outcomes, especially considering the utilisation of multivariable generalised linear models (GLMs), allowing for further insight than previously conducted and highlighting the rigour of the approach in research used. The function of NPI analysis and their effectiveness in the mitigation of certain disease is proved rather most useful in control of spread rather than direct suppression [33, 50], with the use of focusing on timely, stringent implementation paramount to maximising efficacy [33,51]. With the ability to associate NPIs to specific disease, the study distinguishes how NPI implementation may be tiered in importance as similar studies do corroborate the results finding stay-at-home and mass gathering policies as well as most specifically face-coverings being most effective in reducing risk ratios and effective reproduction value [52,53]. The long-term observation was able to support the clear statement, that increasing stringency of NPIs is found to reduce rates of disease with their subsequent relaxation historically resulting in a resurgence of cases [35,53], strongly in respiratory disease [33], with gastrointestinal, vector- and sex-bourne diseases showing varying influence, although high-magnitude correlations when related [54–56].

This study has several limitations that must be acknowledged. Firstly, the bi-weekly data that was utilised, sourced from the Polish Information System for Disease Control and Prevention, inherently carries a bias. This bias stems from various factors, including potential discrepancies in reporting practices and the inability to fully account for individual health perceptions and healthcare-seeking behaviours, which can significantly influence the subsequent diagnosis and reporting of notifiable diseases. Analysis was based on national-level bi-weekly and monthly data, rather than data specific to provinces or cities, which may overlook the spatial heterogeneity of disease patterns within Poland. While the study focused on nine indicators of Non-Pharmaceutical Interventions (NPIs), numerous other variables, such as population vaccination rates, climate variations, and viral mutations, could also impact the prevalence of notifiable diseases. With the extensive data timeline, advances in disease detection would logically increase reported cases, an effect that cannot be accounted for, thus exhibiting a surveillance bias. The complex interplay between NPIs and their outcomes presents a challenge, creating difficulty in independently assessing the precise relationship between each NPI and its corresponding impact on disease outcomes. Another significant assumption in studying the effects of NPIs is of unequivocal adherence. This oversimplifies human behaviour, leading to inaccurate assessments of their effectiveness as adherence varies significantly due to factors like compliance fatigue, cultural differences, and individual risk perceptions. Variability can result in misinterpreting the efficacy of NPIs, particularly when uneven adherence reduces their intended impact or creates unintended consequences, such as false sense of security or inadequate protection. Limitations in testing were also critical, as regions with limited access to COVID-19 tests or other diagnostic tools could not accurately capture infection rates, creating gaps in data and making it difficult to compare regions with different testing capabilities. Moreover, reduced access to medical care and testing for infectious diseases during the early stages of the pandemic and subsequent surges may have obscured the true incidence of cases during those periods. These limitations emphasise the importance of careful interpretation of the findings and consequently highlight an area for future research to further elucidate the effects of NPIs on infectious disease dynamics. Nonetheless, NPIs cannot be treated as isolated features and likely would not have been effective in isolation when compared to their compounding influence on public health. NPIs should not be viewed as standalone measures, as their effectiveness is likely enhanced when considered in conjunction with their broader impact on public health.

Future research should consider more comprehensive methodologies by employing standardised daily data collection techniques that incorporate larger sample sizes from diverse geographic regions and demographic groups. The advent of Big Data enables the utilisation of advanced techniques that surpass traditional methods such as X13-ARIMA-SEATS for deseasonalization and correlation analysis. Furthermore, it allows for the inclusion of additional contextual variables, including adherence to NPIs, socioeconomic status, comorbidities, and environmental factors. This inclusion would provide a more nuanced understanding of NPIs effects on infectious diseases dynamics and would allow for specific selection of NPIs, suiting the particular needs of a group.

### Conclusion

The COVID-19 pandemic, with its unprecedented NPIs, has illuminated critical dynamics in disease prevention and control but also posed new challenges. This study underscores the nuanced role that NPIs have in reducing disease incidence, particularly respiratory infections, yet it also highlights the complexities of population-level immunity as NPIs disrupt typical exposure and immunity patterns. Diseases with strong human-to-human transmission vectors responded rapidly to interventions like face coverings, lockdowns, and movement restrictions, showing reductions in incidence during stringent NPI phases. However, as restrictions eased, the resurgence of infections, particularly respiratory and sexually transmitted diseases, underscores the need for adaptable public health measures. Furthermore, the surge in HIV and syphilis following the Russo-Ukrainian war-driven refugee inflow into Poland underscores how migration events, particularly in post-pandemic settings, can exacerbate vulnerabilities in public health infrastructure. Addressing these epidemiological shifts requires sustained, equity-focused healthcare strategies that prioritise testing, treatment, and surveillance. Looking ahead, the potential for “immunity debt”—resulting from prolonged decreased exposure to common pathogens—warrants careful monitoring. Given the prolonged changes in exposure, as well as new disease dynamics influenced by NPI-induced modifications to the immune landscape, future public health strategies in Poland and similar settings must incorporate agile, evidence-based approaches.

## Data Availability

All data produced in the present study are available upon reasonable request to the authors.

https://www.pzh.gov.pl/

---

**Figure 1.**
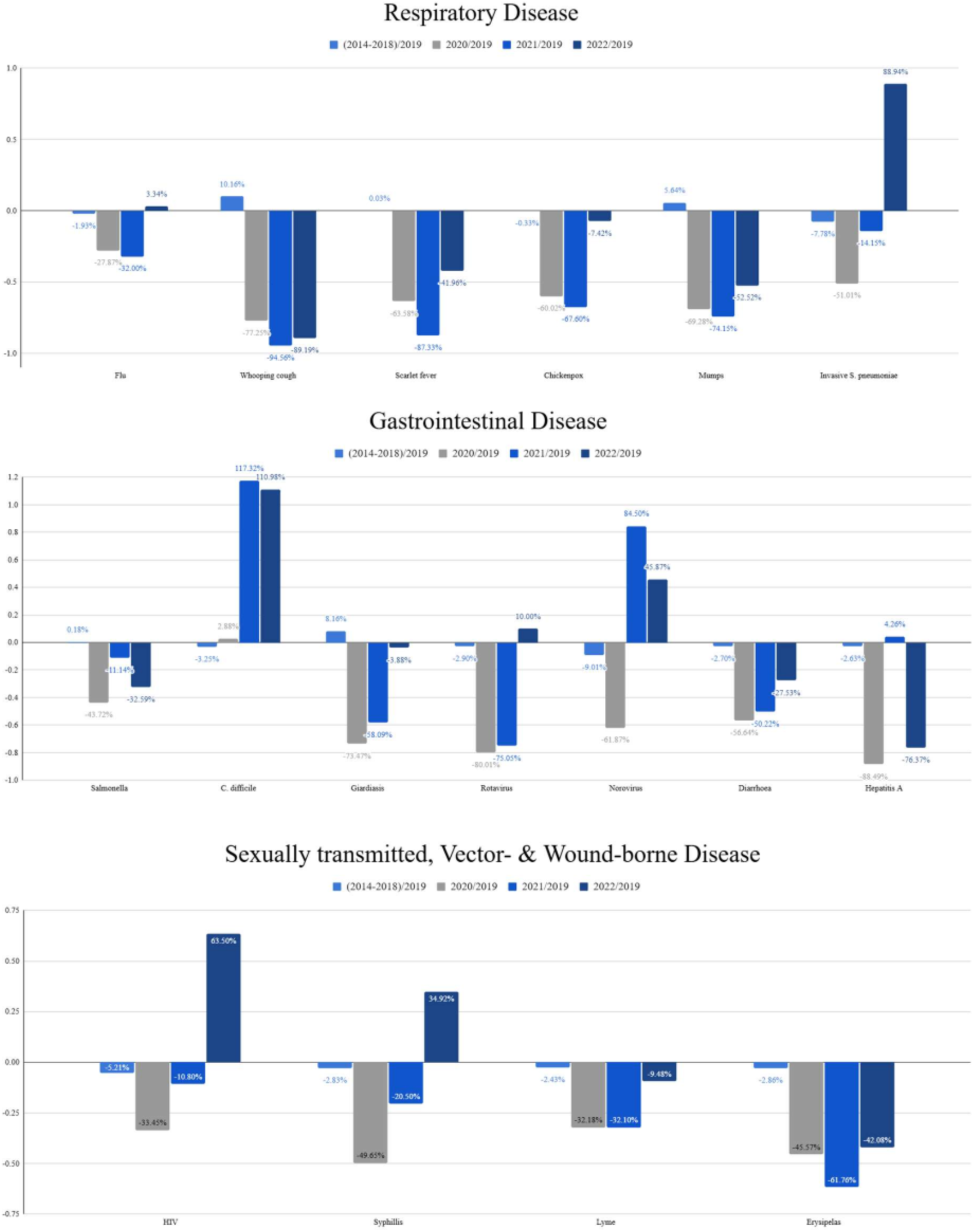
Percentage change of the crude annual incidence of 17 selected notifiable diseases in Poland, before and after the pandemic. (a) Respiratory and air-droplet transmitted diseases, (b) gastrointestinal diseases, (c) Sexually transmitted, vector- and wound-borne diseases.

